# A machine learning-based holistic approach for diagnoses within the Alzheimer’s disease spectrum

**DOI:** 10.1101/2020.10.02.20205559

**Authors:** Noemi Massetti, Alberto Granzotto, Manuela Bomba, Stefano Delli Pizzi, Alessandra Mosca, Reinhold Scherer, Marco Onofrj, Stefano L. Sensi, for the Alzheimer’s Disease Neuroimaging Initiative (ADNI)

## Abstract

Alzheimer’s disease (AD) is a neurodegenerative condition driven by a multifactorial etiology. We employed a machine learning (ML) based algorithm and the wealth of information offered by the Alzheimer’s Disease Neuroimaging Initiative (ADNI) database to investigate the relative contribution of clinically relevant factors for identifying subjects affected by Mild Cognitive Impairment (MCI), a transitional status between healthy aging and dementia. Our ML-based Random Forest (RF) algorithm did not help predict clinical outcomes and the AD conversion of MCI subjects. On the other hand, non-converting (ncMCI) subjects were correctly classified and predicted. Two neuropsychological tests, the FAQ and ADAS13, were the most relevant features used for the classification and prediction of younger, under 70, ncMCI subjects. Structural MRI data combined with systemic parameters and the cardiovascular status were instead the most critical factors for the classification of over 70 ncMCI subjects. Our results support the notion that AD is not an organ-specific condition and results from pathological processes inside and outside the Central Nervous System.

## Introduction

Alzheimer’s disease (AD) is one of the most prevalent forms of dementia (Prince et al., 2013). Clinical and epidemiological evidence indicates that pathogenic changes occur decades before the onset of clinical symptoms (Vermunt et al., 2019). Mild Cognitive Impairment (MCI) is a transitional state between healthy brain aging and dementia and a critical prodromal phase of AD (Morris et al., 2001) that offers a window of opportunity for therapeutic intervention. To date, no disease-modifying strategies are available, and almost all the AD clinical trials have failed (Long and Holtzman, 2019). On the other hand, a growing body of evidence indicates that prevention strategies delay AD onset and progression (Rakesh et al., 2017). Therefore, the development of cost-effective approaches set to identify MCI subjects at-risk of conversion to dementia is of paramount importance.

The clinical identification of the MCI stage occurs through the combined implementation of neuropsychological tests (Jessen et al., 2014; Petersen et al., 1999), the use of brain Magnetic Resonance Imaging (MRI) scans, and the evaluation, in the cerebrospinal fluid (CSF), of altered levels of AD-related proteins (i.e., β-amyloid and tau). Furthermore, experimental and pathological evidence indicates that subtle molecular, cellular, and subcellular changes precede, by decades, the appearance of cognitive symptoms (Dickerson et al., 2009; Scahill et al., 2002; Thompson et al., 2001). Thus, the investigation of these combined biomarkers is critical to set the course of early diagnosis and, in the future, effective therapy.

Machine learning (ML) is a computer science field that provides computational tools that help with automated data classification and event prediction. Applied to AD, ML can capture the complex molecular interactions of pathogenic events that occur in the early stages of the disease and facilitate its progression (Beam and Kohane, 2018; Kononenko, 2001). In that respect, various ML architectures fed with MRI data informative on undergoing subtle structural brain changes have helped generate models of the disease continuum that spans from MCI to AD (Gaser et al., 2013; Hua et al., 2010; Moradi et al., 2015). However, significant levels of accuracy in predicting the MCI transition to dementia have only been achieved by combining detailed MRI-based measurements of cortical thicknesses and brain volumes, the assessment of alterations of β-amyloid and tau in the CSF, comprehensive neuropsychological and behavioral testing, and the evaluation of AD-related genetic mutations as well as sociodemographic data (Bhagwat et al., 2018; Casanova et al., 2013; Lee et al., 2019; Young et al., 2018).

In general, the use of such an array of biomarkers has been limited to parameters that reflect changes occurring within the Central Nervous System (CNS). However, a paradigm shift in the theoretical construct related to dementia and non-transmissible chronic disease indicates that the implementation of a systems medicine and network-based approach along with the evaluation of peripheral and systemic changes may offer better opportunities for the comprehension of the AD spectrum (Lee and Loscalzo, 2019). In line with this network- and systems medicine-based perspective, some authors have proposed that chronic diseases, including dementia, should be considered as the result of converging perturbations of complex intra- and inter-cellular networks and alterations that occur at many levels and not limited to one organ or driven by a single molecular factor or pathogenic mechanism (Barabási et al., 2011; Hampel et al., 2018; Lee and Loscalzo, 2019; Orešič et al., 2010).

Moving from this conceptual mainframe, we have therefore employed an ML-based approach to identify subsets of MCI individuals who are more prone to convert to dementia by taking advantage of a wealth of biological, functional, and MRI-based structural data that reflect the pathogenic events occurring inside as well as outside of the CNS.

To that aim, the study evaluated a cohort of 517 MCI patients and the related data obtained from the Alzheimer’s Disease Neuroimaging Initiative (ADNI) database. In this study cohort, we implemented an ML-based Random Forest (RF) algorithm to classify, in a supervised manner, converting to AD (cMCI) or stable, non-converting (ncMCI), MCI subjects. Data related to the study population were analyzed by RF as separate features or combined and assessed in terms of classification power. In detail, the study investigated the relative contribution in the classification process offered by the use of first-tier diagnostic tools, like alterations of AD-related CSF proteins and neuropsychological tests, or the implementation of second-tier biomarkers, like detailed structural MRI-related brain changes or variations of peripheral parameters. After these first learning steps, the ML-based RF that generated the best classification of cMCI and ncMCI subjects was tested on a new cohort of patients (a test-dataset) to estimate the machine predictive capability. Finally, we performed an RF dividing the MCI study cohort into subjects older or younger than 70. The decision stemmed from the assumption that disease processes occur with a different degree of relevance and pathogenic weight in “younger” versus “older” individuals. The choice was also based on the hypothesis that the pathogenic processes are different and only partially overlapping in these two groups. Thus, in a nutshell, we tested the idea that younger MCI individuals are mainly subjected to primary neurodegenerative processes while older MCI subjects may suffer from a more combined variety of precipitating and age-dependent factors like alterations of blood pressure, energy deficits, and dysmetabolism that also occur outside of the CNS.

## Results

The demographic data of the individuals involved in the study are summarized in Table 1.

**Table 1.** Demographics. The table illustrates the demographics of the MCI cohort (t0) as well as the demographics of the same subjects in the 24-month, for MCI converting to AD (cMCI, t1), and in the 36-month follow-up for MCI subjects non converting do AD (ncMCI, t2).

| | **MCI (n=520)**<br>Time: $t_0$ | **cMCI (n= 178)**<br>Time: $t_1$ | **ncMCI (n= 342)**<br>Time: $t_2$ |
| --- | --- | --- | --- |
| **Gender (female/male)** | 196 / 325 | 70 / 108 | 126 / 216 |
| **Age (years)** | 73.6 ± 7.6 | 75.2 ± 7.3 | 76.4 ± 7.8 |
| **education (y)** | 15.8 ± 2.9 | 15.8 ± 2.7 | 15.8 ± 2.9 |
| **MMSE** | 27.3 ± 1.9 | 23.7 ± 2.9 | 27.1 ± 2.9 |
| **ADAS13** | 17.7 ± 6.9 | 26.9 ± 6.8 | 16.6 ± 8.8 |
| **ApoE ε4 (Non carrier/Het/Homo)** | 252 / 207 / 61 | 57 / 91 / 30 | 195 / 116 / 31 |

The best combinations of features that allow the most accurate RF-driven classification of cMCI and ncMCI patients, as well as their performance evaluation, are shown in Table 2. Results are depicted and sorted by accuracy values. Predictivity values were considered significant if greater than or equal to 85%. Results of the whole ML analysis are shown in Supplementary Tables 1 and 2. In cMCI patients, no combination of parameters allowed the ML to achieve a significant predictivity value (Table 2). Thus, disappointingly, we had to conclude that ML could not detect cMCI subjects when the machine is only fed with baseline data of these individuals.

**Table 2.** ML prediction performance for MCI subjects converting (cMCI) and non-converting (ncMCI) to AD. The tables depict the ML ability to classify cMCI and ncMCI subjects in the test dataset. The ranking is based on Accuracy values.

|  | cMCI |  |  |  |  |  | ncMCI |  |  |  |  |
| --- | --- | --- | --- | --- | --- | --- | --- | --- | --- | --- | --- |
|  | Number of trees | Predictivity | Accuracy | Sensitivity | Specificity |  | Number of trees | Predictivity | Accuracy | Sensitivity | Specificity |
| NEURO | 500 | 66.8% | 0.95 | 0.87 | 1.00 | NEURO | 500 | 84.9% | 0.95 | 1.00 | 0.87 |
| NEURO+EDU | 300 | 66.2% | 0.95 | 0.87 | 1.00 | NEURO+EDU | 300 | 86.4% | 0.95 | 1.00 | 0.87 |
| TRI_COL | 300 | 12.70% | 0.92 | 0.83 | 0.97 | TRI_COL | 300 | 87% | 0.92 | 0.97 | 0.83 |
| ENER_SUB | 250 | 22.70% | 0.92 | 0.79 | 1.00 | ENER_SUB | 250 | 84.70% | 0.92 | 1.00 | 0.79 |
| SYST_IND | 200 | 35.40% | 0.92 | 0.83 | 0.97 | SYST_IND | 200 | 74.10% | 0.92 | 0.97 | 0.83 |
| AMINO | 150 | 31.20% | 0.90 | 0.79 | 0.97 | AMINO | 150 | 72.90% | 0.90 | 0.97 | 0.79 |
| NEURO+METABOL | 200 | 59.6% | 0.86 | 0.86 | 0.87 | NEURO+METABOL | 200 | 85.7% | 0.86 | 0.87 | 0.86 |
| CSF+ENER_SUB | 300 | 44.70% | 0.86 | 1.00 | 0.75 | CSF+ENER_SUB | 300 | 82.10% | 0.86 | 0.75 | 1.00 |
| CSF+VIT_SIG | 250 | 55.5% | 0.82 | 0.64 | 0.83 | CSF+VIT_SIG | 250 | 77.3% | 0.82 | 0.83 | 0.64 |
| NEURO+VIT_SIG | 300 | 59.6% | 0.81 | 0.54 | 0.93 | NEURO+VIT_SIG | 300 | 84.2% | 0.81 | 0.93 | 0.54 |
| NEURO+ApoEε4 | 300 | 64.9% | 0.81 | 0.60 | 0.94 | NEURO+ApoEε4 | 300 | 89.2% | 0.81 | 0.94 | 0.60 |
| MRI+BIL_ACID | 300 | 51.7% | 0.80 | 0.75 | 0.84 | MRI+BIL_ACID | 300 | 89.9% | 0.80 | 0.84 | 0.75 |
| MRI+CSF | 250 | 58.9% | 0.78 | 0.64 | 0.85 | MRI+CSF | 250 | 86.7% | 0.78 | 0.85 | 0.64 |
| MRI+NEURO+CSF | 250 | 69.4% | 0.78 | 0.64 | 0.85 | MRI+NEURO+CSF | 250 | 88.6% | 0.78 | 0.85 | 0.64 |
| CSF+AMINO | 300 | 52% | 0.78 | 0.68 | 0.81 | CSF+AMINO | 300 | 79.20% | 0.78 | 0.81 | 0.68 |
| NEURO+BIL_ACID | 300 | 60.4% | 0.77 | 0.60 | 0.86 | NEURO+BIL_ACID | 300 | 87.0% | 0.77 | 0.86 | 0.60 |
| NEURO+MRI | 300 | 61.1% | 0.75 | 0.56 | 0.87 | NEURO+MRI | 300 | 89.0% | 0.75 | 0.87 | 0.56 |

### The ML-based RF helps in the classification of ncMCI subjects

The ML performance in classifying ncMCI subjects was very effective. When employing data related to the neuropsychological tests (NEURO), we reached an encouraging accuracy value of 0.95 and a predictivity rate of 84.9% (Table 2). The result improved and reached a predictivity value of 86.4% when education levels were added (Table 2). Interestingly, the neuropsychological test that had a decisive weight for the classification was the Rey Auditory Verbal Learning Test (RAVLT) immediate, a test employed to assess episodic memory. However, to our surprise, data related to biological samples generated the best predictivity value (Table 2). Triglyceride and cholesterol (TRI_COL) features allowed the RF to correctly predict 87% of the ncMCI individuals with an accuracy rate of 0.92. The same accuracy value was reached when feeding the ML with features related to energy substrates (ENER_SUB) and systemic indices like changes in blood levels of albumin and creatinine (SYST_IND), even though only in the first two cases the predictivity approached the threshold for statistical significance and produced a predictivity value of 84.7%. Of note, the most determinant features for the classification and prediction were changes related to blood levels of triglycerides, low-density lipoproteins (LDL), and glucose.

### The ML-based RF analysis reveals at least two different undergoing conditions and the presence of an age-divide

To investigate the effect of age, we divided the study cohort into cMCI subjects over and under 70 years of age. Table 3 shows the best results for the two cMCI groups sorted by considering the ML-based RF performance accuracy. No combination of data allowed the ML to classify and predict the fate of cMCI patients.

**Table 3.** ML prediction performance for MCI subjects converting to AD (cMCI), following age stratification. The tables depict the ML ability to classify subjects following age stratification as indicated. Measurements of predictivity, accuracy, sensitivity, and specificity refer to the ML performance obtained from the test-dataset. Please, note that, in both cases, the predictivity values do not reach the 85% threshold value. The ranking is based on the accuracy score.

|  | cMCI (> 70) |  |  |  |  |
| --- | --- | --- | --- | --- | --- |
|  | Number of trees | Predictivity | Accuracy | Sensitivity | Specificity |
| MRI+METABOL | 300 | 54.3% | 0.98 | 0.95 | 1.00 |
| MRI+VIT_SIGN | 300 | 52.4% | 0.98 | 0.95 | 1.00 |
| MRI+SYST_IND | 350 | 53% | 0.95 | 0.90 | 1.00 |
| TRI_COL | 250 | 24.80% | 0.93 | 0.85 | 1.00 |
| ENER_SUB | 250 | 25.50% | 0.93 | 0.85 | 1.00 |
| AMINO | 200 | 36.20% | 0.93 | 0.85 | 1.00 |
| NEURO | 500 | 59.0% | 0.89 | 0.84 | 0.91 |
| NEURO+EDU | 200 | 61.8% | 0.89 | 0.84 | 0.91 |
| SYST_IND | 250 | 37.10% | 0.88 | 0.85 | 0.91 |
| NEURO+CSF | 300 | 76.0% | 0.88 | 0.70 | 0.96 |
| NEURO+CSF+ApoEε4 | 200 | 73.3% | 0.88 | 0.80 | 0.92 |
| NEURO+ApoEε4 | 350 | 65.4% | 0.87 | 0.79 | 0.91 |
| CSF+VIT_SIGN | 250 | 54.8% | 0.83 | 0.75 | 0.87 |
| NEURO+VIT_SIGN | 400 | 52.3% | 0.81 | 0.65 | 0.91 |
| NEURO+ENER_SUB | 200 | 59% | 0.78 | 0.55 | 0.94 |
| NEURO+BIL_ACID | 300 | 57.1% | 0.77 | 0.50 | 0.89 |
| CSF+METABOL | 300 | 49.3% | 0.76 | 0.56 | 0.83 |

|  | cMCI (< 70) |  |  |  |  |
| --- | --- | --- | --- | --- | --- |
|  | Number of trees | Predictivity | Accuracy | Sensitivity | Specificity |
| NEURO | 400 | 67.4% | 1.00 | 1.00 | 1.00 |
| NEURO+EDU | 200 | 71.4% | 1.00 | 1.00 | 1.00 |
| NEURO+MRI | 350 | 63.8% | 0.95 | 1.00 | 0.93 |
| MRI+NEURO+CSF | 300 | 65.4% | 0.93 | 0.50 | 1.00 |
| MRI | 400 | 52.7% | 0.90 | 0.67 | 1.00 |
| CSF+VIT_SIGN | 350 | 31.0% | 0.88 | 1.00 | 0.86 |
| CSF+SYST_IND | 200 | 57.10% | 0.88 | 0.00 | 1.00 |
| CSF+AMINO | 200 | 53.60% | 0.88 | 0.00 | 1.00 |
| MRI+CSF | 250 | 57.7% | 0.86 | 0.00 | 1.00 |
| NEURO+VIT_SIGN | 300 | 72.5% | 0.78 | 0.43 | 0.94 |
| NEURO+CSF | 300 | 64.0% | 0.76 | 0.63 | 0.89 |
| NEURO+CSF+ApoEε4 | 200 | 68.0% | 0.76 | 0.63 | 0.89 |
| TRI_COL | 250 | 15.60% | 0.73 | 0.00 | 1.00 |
| ENER_SUB | 250 | 21.80% | 0.73 | 0.00 | 1.00 |
| SYST_IND | 250 | 21.80% | 0.73 | 0.00 | 1.00 |
| AMINO | 200 | 25% | 0.73 | 0.00 | 1.00 |
| MRI+ENER_SUB | 400 | 53.10% | 0.73 | 0.50 | 0.82 |

The situation was different when investigating younger or older ncMCI individuals. The best RF’s predictivity values increased, and, interestingly, the data combination that helped the better ML performance completely differed (Table 4). In older ncMCI individuals, structural MRI data combined with features related to lipoproteins, glucogenesis-associated metabolites, and amino acids (METABOL) allowed the best classification and prediction model generating the highest values of accuracy and predictivity (Supplementary Figure 1). The left hippocampus volume was the MRI-related feature that generated a prediction value of 85.5% and an accuracy of 0.98. Feeding the RF with MRI data associated with vital signs (VIT_SIG) produced the second-best result, although the predictivity value dropped to 82.1%. Finally, combining MRI data with systemic indices (SYST_IND) allowed the RF to classify ncMCI subjects with an accuracy value of 0.95 and a predictivity value of 86.3% (Table 4).

**Table 4.** ML prediction performance for MCI subjects non-converting to AD (ncMCI), following age stratification. The tables depict the ML ability to classify subjects following age stratification as indicated. Measurements of predictivity, accuracy, sensitivity, and specificity refer to the ML performance obtained from the test-dataset. Please, note that, unlike cMCI (Table 3), the predictivity value exceeds the 85% threshold, with ML performance for ncMCI subjects < 70 years showing the highest predictivity scores. The ranking is based on the accuracy score.

|  | ncMCI (> 70) |  |  |  |  |
| --- | --- | --- | --- | --- | --- |
|  | Number of trees | Predictivity | Accuracy | Sensitivity | Specificity |
| MRI+METABOL | 300 | 85.5% | 0.98 | 1.00 | 0.95 |
| MRI+VIT_SIGN | 300 | 82.1% | 0.98 | 1.00 | 0.95 |
| MRI+SYST_IND | 350 | 86.30% | 0.95 | 1.00 | 0.90 |
| TRI_COL | 250 | 81.40% | 0.93 | 1.00 | 0.85 |
| ENER_SUB | 250 | 81.90% | 0.93 | 1.00 | 0.85 |
| AMINO | 200 | 73.90% | 0.93 | 1.00 | 0.85 |
| NEURO | 500 | 84.6% | 0.89 | 0.91 | 0.84 |
| NEURO+EDU | 200 | 85.1% | 0.89 | 0.91 | 0.84 |
| SYST_IND | 250 | 77.10% | 0.88 | 0.91 | 0.85 |
| NEURO+CSF | 300 | 81.8% | 0.88 | 0.96 | 0.70 |
| NEURO+CSF+ApoEε4 | 200 | 80.1% | 0.88 | 0.92 | 0.80 |
| NEURO+ApoEε4 | 350 | 86.6% | 0.87 | 0.91 | 0.79 |
| CSF+VIT_SIGN | 250 | 74.4% | 0.83 | 0.87 | 0.75 |
| NEURO+VIT_SIGN | 400 | 85.0% | 0.81 | 0.91 | 0.65 |
| NEURO+BIL_ACID | 300 | 87.0% | 0.77 | 0.89 | 0.50 |
| CSF+METABOL | 300 | 76.9% | 0.76 | 0.83 | 0.56 |
| MRI+NEURO+CSF | 300 | 87.8% | 0.74 | 0.88 | 0.50 |

|  | ncMCI (< 70) |  |  |  |  |
| --- | --- | --- | --- | --- | --- |
|  | Number of trees | Predictivity | Accuracy | Sensitivity | Specificity |
| NEURO | 400 | 90.2% | 1.00 | 1.00 | 1.00 |
| NEURO+EDU | 200 | 88.0% | 1.00 | 1.00 | 1.00 |
| NEURO+MRI | 350 | 86.0% | 0.95 | 0.93 | 1.00 |
| MRI+NEURO+CSF | 300 | 89.5% | 0.93 | 1.00 | 0.50 |
| MRI | 400 | 87.3% | 0.90 | 1.00 | 0.67 |
| CSF+VIT_SIGN | 350 | 85.5% | 0.88 | 0.86 | 1.00 |
| CSF+SYST_IND | 200 | 87.90% | 0.88 | 1.00 | 0.00 |
| CSF+AMINO | 200 | 89.40% | 0.88 | 1.00 | 0.00 |
| MRI+CSF | 250 | 89.5% | 0.86 | 1.00 | 0.00 |
| NEURO+VIT_SIGN | 300 | 90.0% | 0.78 | 0.94 | 0.43 |
| NEURO+CSF | 300 | 94.6% | 0.76 | 0.89 | 0.63 |
| NEURO+CSF+ApoEε4 | 200 | 91.9% | 0.76 | 0.89 | 0.63 |
| TRI_COL | 250 | 91% | 0.73 | 1.00 | 0.00 |
| ENER_SUB | 250 | 93.30% | 0.73 | 1.00 | 0 |
| SYST_IND | 250 | 86.60% | 0.73 | 1.00 | 0 |
| AMINO | 200 | 86.60% | 0.73 | 1.00 | 0 |
| MRI+TRI_COL | 300 | 12.80% | 0.73 | 0.82 | 0.50 |

For younger ncMCI subjects, the scenario was different. Data related to neuropsychological scores alone allowed the RF to correctly classify with a 100% accuracy and 90.2% predictivity. These values were achieved by the interaction of all the employed 16 neuropsychological tests. Of note, the use of the Functional Activity Questionnaire (FAQ; a questionnaire used for the assessment of daily living activities) and the Alzheimer’s Disease Assessment Scale–Cognitive Subscale test (ADAS13; a test that assesses the severity of impairment to memory, learning, language, practice, and orientation), as well as the RAVLT-immediate test, were the primary driving force for the RF process (Supplementary Figure 2). Combining neuropsychological evaluations with education levels decreased the predictivity to 88%, although the accuracy score was unaffected. In contrast, the combination of neuropsychological test scores with MRI structural data was less useful to help the ML in the classification and produced an accuracy value of 0.95 and a prediction score of 86%.

Results of our ML-based RF approach indicate that the tool cannot help predict the clinical outcomes of cMCI subjects. On the other hand, ncMCI individuals were correctly classified and predicted. In particular, our results indicate that two behavioral/neuropsychological tools like the FAQ and ADAS13 are the most relevant for the classification and prediction of younger ncMCI subjects. At the same time, structural MRI data combined with data informative on the status of energy and lipid metabolisms as well as measurements of patient’s cardiovascular status and systemic indices were the most critical factors for the classification and prediction of older ncMCI individuals.

## Discussion

In this study, we aimed at identifying which combination of ADNI-related data was the most effective for the classification of cMCI and ncMCI patients. We started with CNS-related features that are classically employed in the dementia diagnostic workup to ascertain neurodegeneration and cognitive decline (i.e., altered levels of AD-related CSF proteins and neuropsychological evaluations). We then added structural MRI features and peripheral biomarkers (i.e., blood/plasma levels of lipoproteins, gluconeogenic metabolites, amino acids, bile acids, and purines) to refine the analyses. This large number of variables, assessed only at baseline, was not enough for the ML to correctly classify and predict the fate of cMCI subjects. The RF failed to achieve significant accuracy and predictivity when estimating the conversion in young and old MCI individuals. One possible explanation for the poor performance is that the ML-based RF needs more than baseline features to work, thereby making the availability of data indicative of longitudinal changes mandatory.

A very different scenario emerged when approaching the ncMCI dataset. In this case, baseline data were sufficient to classify and identify stable MCI patients with significant accuracy levels. The analysis of the features employed to allocate these subjects indicates that neuropsychological tests, triglycerides, cholesterol, and energy substrates were the major driving forces for the RF process. This result supports the hypothesis that AD, like other chronic diseases, is not an organ-specific condition but the result of a wealth of mostly unexplored pathological processes that occur inside and outside of the CNS. Further inferences were possible when dividing ncMCI individuals upon the 70-year-old threshold.

Older ncMCI subjects were best classified by using a combination of detailed structural MRI and lipid- and energy metabolism-related features (METABOL), cardiovascular measurements (VIT_SIGN), along with systemic indices (SYST_IND), thereby indicating the convergence of pathogenic processes that are likely occurring inside and outside of the brain (Fig. 1). These findings are in line with previous studies showing the occurrence of lipid and energy-related dysmetabolism in AD patients (Bernath et al., 2020; Butterfield and Halliwell, 2019; Cunnane et al., 2020; Gamberger et al., 2017). The RF output analysis indicated that features related to left hippocampal volumes had the most weight in the classification, in line with well-established observations indicating that this region is the most affected by the AD-related neurodegenerative processes (Apostolova et al., 2006; Fox et al., 1996; Henneman et al., 2009).

On the other hand, when assessing young ncMCI individuals, the best prediction and classification performances were achieved using cognitive status features (Fig. 1). The items that showed more classification power were the FAQ and the ADAS13. This finding is not surprising as the FAQ is an ecological tool used to assess the subject’s ability to deal with daily activities and personal care. These features are deeply affected by aging but expected to be less defective in younger patients and, therefore, if impaired, interpreted as significant warning signs by the ML.

The ADAS13 evaluates the overall cognitive status (Kueper et al., 2018). Of note, in routine clinical work, the MMSE is preferred to the ADAS13, but, surprisingly, our ML-based RF found the MMSE scores almost the least useful parameter for these patients’ classification. This result can be explained by considering the origin and score structure of the two tests.

The MMSE was created as an easy-to-use clinical tool, while the ADAS13 is more research-oriented (Kueper et al., 2018). The score range is also different, more comprehensive [from 0 (no symptoms) to 70 (maximum symptoms)] in the ADAS13 compared to the limited MMSE 30 points (with 30 indicative of no symptoms and 0 of maximum symptoms). Thus, the ADAS13 is more specific and offers a more granular scale of values that can assess more subtle cognitive abnormalities (Schmidt-Richberg et al., 2016).

Finally, early cognitive and memory deficits are signs of undergoing synaptic failure (DeKosky and Scheff, 1990; Selkoe, 2002; Sheng et al., 2012). However, these changes are behind the resolution power of clinical MRI protocols, which might explain why our RF made little use of MRI features to classify younger patients. On the other hand, in older individuals, synaptic alterations vanish and dilute in the context of a broader degeneration detected by MRI, thereby allowing a better ML classification performance of ncMCI subjects.

## Conclusions

AD is a complex and multifactorial condition, and the characterization of patients in a prodromal stage of the disease (MCI) represents a challenge for biomedical research and unmet clinical and therapeutic needs.

A revision of the dementia construct may help to reach these targets. For instance, a growing body of evidence supports the view that AD, Parkinson’s disease (PD), and Amyotrophic Lateral Sclerosis (ALS), to name the most frequent neurodegenerative conditions, share key pathophysiological processes (Espay et al., 2019). Clinical and epidemiological evidence have indicated that the AD-, PD- and ALS-related neurodegenerative processes result from the converging activity of many polygenic, epigenetic, environmental, vascular, excitotoxic, and metabolic factors (Brem and Sensi, 2018; Sensi et al., 2018). In the case of AD, a growing body of evidence supports the notion that many patients exhibit mixed neuropathology and different arrays of neurotoxic protein aggregates like Aβ, Tau, prion proteins (PrP), α-synuclein, and TAR DNA binding protein-43 (TDP-43) (Boyle et al., 2018).

In the past thirty years, a monumental effort in financial and human resources has been made to reduce these aggregated proteins. The rationale behind this strategy is that protein deposits are “toxic” and their physical disaggregation could stop neurodegeneration progression (Mathieu et al., 2020). Except for few highly debated clinical trials, the strategy has failed, thereby casting some fundamental doubts on the construct’s validity (Cummings et al., 2017; Herrup, 2015; Knopman et al., 2020; Morris et al., 2018).

ML techniques and big data analysis can help identify novel and unexpected disease features and escape the dogmatic loop in which we are currently entrapped. For instance, a surprising finding of our study concerns the importance of peripheral biomarkers. The analysis of ML variables employed for classifying older ncMCI subjects revealed that - along with neuropsychological tests and changes in blood levels of triglycerides, cholesterol, and energy substrates - one of the most discriminating features used by our RF were the variations of blood levels of bile acids (BA), including the tauroursodeoxycholic acid (TUDCA). This intriguing result is in line with a growing body of evidence supporting the gut-brain connection, the role of the microbiota in neurodegeneration, and the role played by the liver in these processes (Sampson et al., 2016; Zhu et al., 2020). The notion is also supported by a recent study in AD patients indicating the association between altered BA profiles with higher degrees of brain atrophy, brain hypometabolism (as assessed by FDG-PET), and alterations of CSF AD-related biomarkers (Nho et al., 2019). These findings also agree with a study in which AD patients exhibited significantly low plasma levels of several medium-chain acylcarnitines (ACC) (Ciavardelli et al., 2016). These changes indicate hepatic dysfunction as most of the Fatty Acid Oxidation (FAO), the mechanism that controls ACC production (Schooneman et al., 2013), is controlled by the liver.

Of note, defective hepatic FAO eventually impairs ketogenesis and produces lower levels of plasma ketones (Fukao et al., 2004), the brain’s energy substrates alternative to glucose. Thus, the impairment of hepatic ketogenesis occurring in AD may further exacerbate the energetic brain deficits and be a critical aggravating factor for disease progression. Interestingly, in preclinical AD models and cognitively impaired subjects, MCI or AD patients, ketogenic diets and/or pharmacologic manipulations that favor ketogenesis have been shown to improve cognitive performances (Fortier et al., 2020; Henderson et al., 2009; Reger et al., 2004; Van der Auwera et al., 2005; Yao et al., 2011). These AD-related peripheral changes are also in agreement with recent findings indicating that TUDCA, together with sodium phenylbutyrate, can be a promising disease-modifying strategy for ALS (Paganoni et al., 2020).

This set of combined systemic alterations is the gateway to precision medicine and offers fertile ground for innovative research. Most importantly, the search for a cure will likely depend on exploring unchartered territories and/or applying an “outside of the box” line of thinking. Artificial Intelligence and its unbiased computational capability will provide the tools to understand the unique complexity of “individual” diseases affecting each of our patients and take full advantage of a precision medicine-based approach. Precision medicine, systems medicine, and network-based approaches will generate tailored diagnoses, predict disease risks, and produce customized treatments that maximize safety and efficacy (Greene and Loscalzo, 2017; Loscalzo and Barabasi, 2011).

In that vein, we hope that our study can contribute to further exploring ML models to understand the mystery of neurodegenerative processes and dementia.

## Methods

All the data used in this work are available from ADNI database. ADNI is a public-private repository of clinical, imaging, genetic, and biochemical biomarker data obtained from North American subjects or patients (www.adni-info.org). ADNI aims at identifying the determinant processes leading to AD and diagnose pathological changes occurring at the earliest stage. The data downloaded and analyzed in this study were collected at the first appointment (baseline) of the follow-up series required by the ADNI protocols. The management and selection of the data were carried out with custom made R-written codes.

### Subjects

Subjects considered in this study were patients diagnosed with MCI extracted from the cohorts of ADNI-1, ADNI-GO, ADNI-2, and ADNI-3. The inclusion criteria for these patients were the ones provided by the ADNI protocols, and the diagnosis of MCI based on the assessment of global cognition as well as scores of neuropsychological evaluations related to episodic memory, the presence of subjective cognitive disorders, and the clinical evaluation through a semi-structured interview.

Two classes of MCI subjects were chosen: 1) individuals with a baseline diagnosis of MCI who converted to AD within 24 months (cMCI) and individuals with a diagnosis of MCI at baseline who remained stable for the following 36 months (ncMCI). The timeframe was chosen taking into consideration evidence indicating that subjects with a diagnosis of MCI have a high probability of converting to AD within 30 months (Fischer et al., 2007).

### Neuropsychological assessments

For each person, we considered sixteen neuropsychological tests employed to assess the status of different cognitive domains. This neuropsychological dataset included the Alzheimer’s Disease Assessment Scale-Cognitive (ADAS-Cog) subscales to evaluate the severity of memory, learning, language (production and comprehension), praxis, and orientation deficits (Mohs and Cohen, 1988),(Mohs et al., 1997); the Mini-Mental State Examination (Folstein et al., 1975) to investigate global cognition; the 30-item Boston Naming Test (E, Goodglass H, 1983) and the Animal Fluency (Morris et al., 1989) to assess semantic memory and language abilities; the Functional Activities Questionnaire (FAQ) for the assessment of daily living activities (Pfeffer et al., 1982); the Rey Auditory Verbal Learning Test and Logical Memory II, subscale of the Wechsler Memory Scale-Revised (WMS-R) to investigate recall and recognition (Rey, 1964; Wechsler, 1997); the Trail Making Test (Spreen, O., y Strauss, 1998), part A and B (time to completion) to assess attention/executive functions; the Clock Drawing Test to evaluate attention, working and visual memory, and auditory comprehension (Nishiwaki et al., 2004); the Clinical Dementia Rating Scale to quantify the patient degree of dementia (Tractenberg et al., 2001). The Supplementary Table 3 shows the abbreviations used for each neuropsychological assessment and the domains and cognitive functions that each test is set to evaluate.

### Biospecimen quantification

Supplementary Table 4 shows the biomarkers and the biospecimens considered in this work. The biospecimen selection, within the datasets available on the ADNI database (Biospecimen Inventory, http://adni.loni.usc.edu), was made by taking into account the number of samples and the consistency of the biospecimen measurements among the different phases of the ADNI project (ADNI-1, ADNI-GO, ADNI-2, ADNI-3). To meet the second criterion and reduce the incidence of human error, we considered only data produced through automated techniques.

### Additional indices and parameters

ADNI data related to years of education (Stern, 2006), APOE-ε4 genotypes (Blacker et al., 1997), and “vital signs” like systolic (VSBPSYS) and diastolic pressure (VSBPDIA), ventilation frequency (VSRESP), cardiac frequency (VSPULSE), and body temperature (VSTEMP) were also extracted.

### Structural Magnetic Resonance data

The MRI data downloaded from the ADNI database (Image Collections, http://adni.loni.usc.edu) were acquired with a Magnetization Prepared-RApid Gradient Echo (MP-RAGE) protocol by employing a Philips 3-tesla MRI scanner (for details about MRI procedures and protocols see http://adni.loni.usc.edu/wp-content/uploads/2010/05/ADNI2_MRI_Training_Manual_FINAL.pdf) thereby limiting bias and technical issues associated with the use of different scanner types/brand. T1-weighted images were obtained using 3D Turbo Field-Echo sequences (slice thickness of 1.2 mm, repetition time/echo time of 6.8/3.1 ms). The structural MRI analysis was performed with Freesurfer (version 6.0). Automatic reconstruction and labeling of cortical and subcortical regions was achieved with the “recon-all-all” command line, according to Desikan-Killiany Atlas (Desikan et al., 2006). The volumes of the brain regions, computed with *asegstats2table*, were normalized by dividing to the total intracranial volume of each patient, while the thicknesses of the brain areas considered are those calculated automatically by *aparcstats2table*. Supplementary Table 3 shows brain areas analyzed in this study.

### ML analysis

Our ML approach implemented an RF algorithm. The RF is a supervised non-linear classifier, and its operation is based on the construction of binary decision trees obtained with the Bagging sampling method (an acronym for bootstrap aggregating) (Breiman, 1996). The RF classifier was chosen because the performance is robust and stable over an extensive range of parameters. Furthermore, it is independent of the distribution of data (i.e., neuropsychological scores, quantifications of lipoproteins, etc.) and has significant multi-class and advanced data-mining capabilities (Steyrl et al., 2016).

RF analyzed the data obtained from the ADNI baseline. In the training phase, the algorithm studied the non-linear interactions between the data (or features) of the subjects divided upstream into the two classes: cMCI and ncMCI. The goal at this stage was to identify the best subdivision/classification strategy. In the testing phase, the approach was assayed on a cohort of new subjects.

In the training phase, the RF analyzed 85% of each dataset (randomly extracted) and performed 30 times the analysis; the number of trees was chosen manually and set to minimize the out-of-bag error (number of trees: 200, 250, 300, 350, 400, 450, 500, 550, 600).

The parameters considered to evaluate the ML performance in classifying cMCI and ncMCI subjects were *accuracy, sensitivity*, and *specificity*. To assess the ability to predict whether a subject belongs to one of the two classes and, therefore, investigate, separately, the classification strategy for cMCI and ncMCI subjects, we introduced the variable *predictivity*, which measures the percentage of subjects (test-dataset) correctly classified in each of the two classes.

In the study, we employed the Treebagger class function of the Statistics and Machine Learning Toolbox of MATLAB (The Mathworks Inc., Natick, MA, USA).

**Figure 1.**
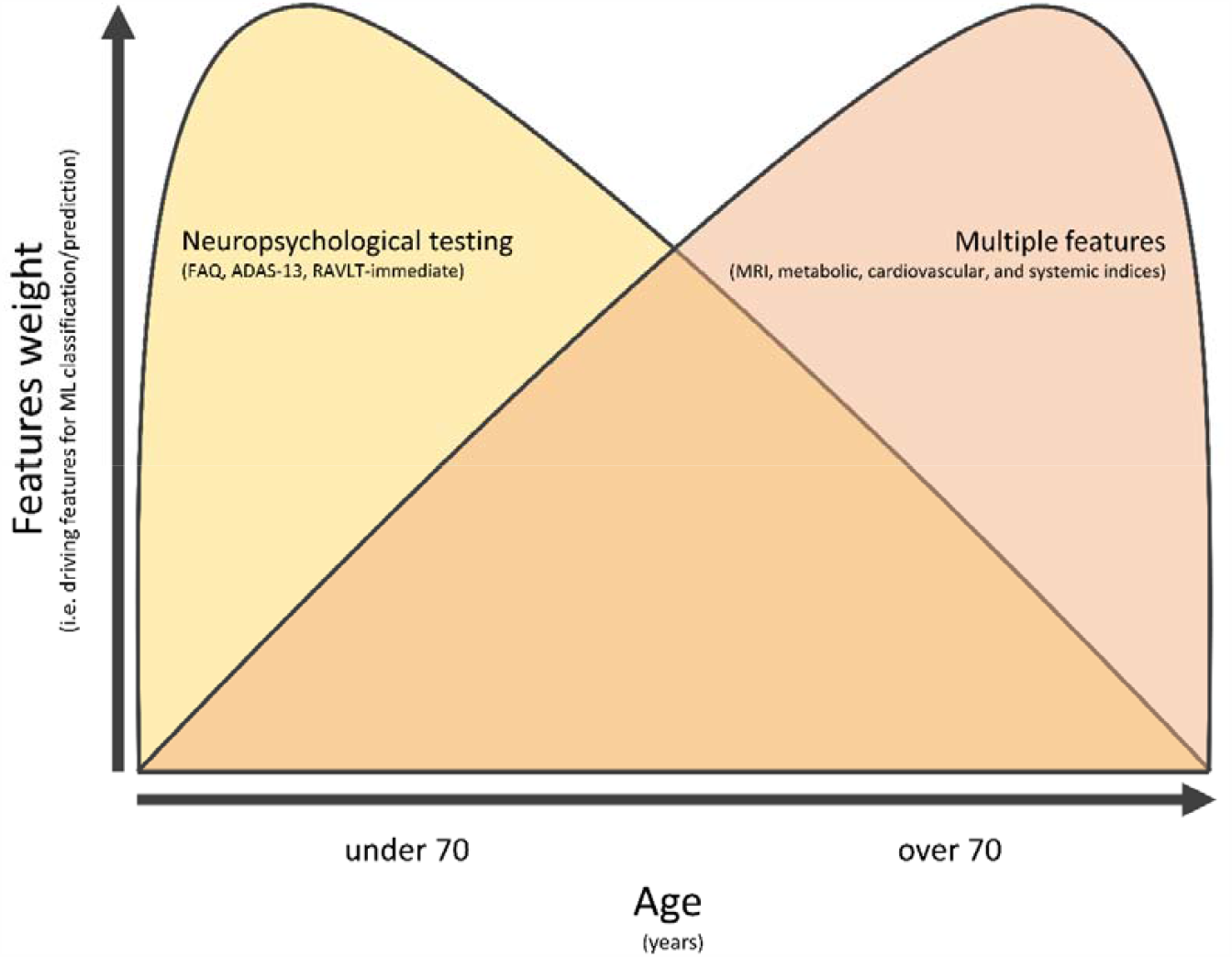
Differential contribution of ADNI-derived features for the classification and prediction of ncMCI. The pictogram illustrates the differential contribution of ADNI-derived features (see Supplementary Information for a complete list of the assayed items) employed by the ML-based RF algorithm to classify and predict ncMCI subjects. Note that the 70-year-old age cutoff reveals a marked dichotomy on how the RF algorithm employs and weights different features to discriminate “younger” and “older” individuals. For subjects < 70 years of age, the classification/prediction is primarily driven by neuropsychological testing (FAQ, ADAS13, RAVLT-immediate). On the contrary, subjects > 70 years of age are identified by a more structured palette of imaging, metabolic, and systemic data.

## Data Availability

The data will be made available upon request.

## Acknowledgments

The study was funded by the Alzheimer’s Association Part the Cloud: Translational Research Funding for Alzheimer’s Disease (PTC) PTC-19-602325 and the Alzheimer’s Association - GAAIN Exploration to Evaluate Novel Alzheimer’s Queries (GEENA-Q-19-596282) (SLS). AG is supported by the European Union’s Horizon 2020 Research and Innovation Program under the Marie Sklodowska-Curie grant agreement iMIND—No. 841665.

## Conflict of interests

None of the authors have conflict of interests. This study is not industry-sponsored.

## Supplementary information

The manuscript includes the following supplementary material:

Supplementary File 1

Supplementary File 2

Supplementary File 3

Supplementary File 4

Supplementary File 5

Supplementary File 6

Supplementary File 7

## Notes

### Competing Interest Statement

The authors have declared no competing interest.

### Author Declarations

Data were obtained upon permission from the public ADNI database (last protocol update 10/17/2019). The ADNI study is conducted according to Good Clinical Practice guidelines, US 21CFR Part 50 Protection of Human Subjects, and Part 56 Institutional Review Boards(IRBs) / Research Ethics Boards (REBs), and pursuant to state and federal HIPAA regulations. Phone consents was obtained for all prescreening procedures and written informed consent for the study was obtained from all participants and/or authorized representatives and the study partners before in person assessments were carried out (in accordance with US 21 CFR 50.25, the Tri Council Policy Statement). All the procedures employed in the ADNI study are in compliance with national and international regulations.

