## Supplementary Table 3 for "A machine learning-based holistic approach for diagnoses within the Alzheimer’s disease spectrum"

**Supplementary Table 3.** Abbreviations of neuropsychological tests and the associated cognitive domains.

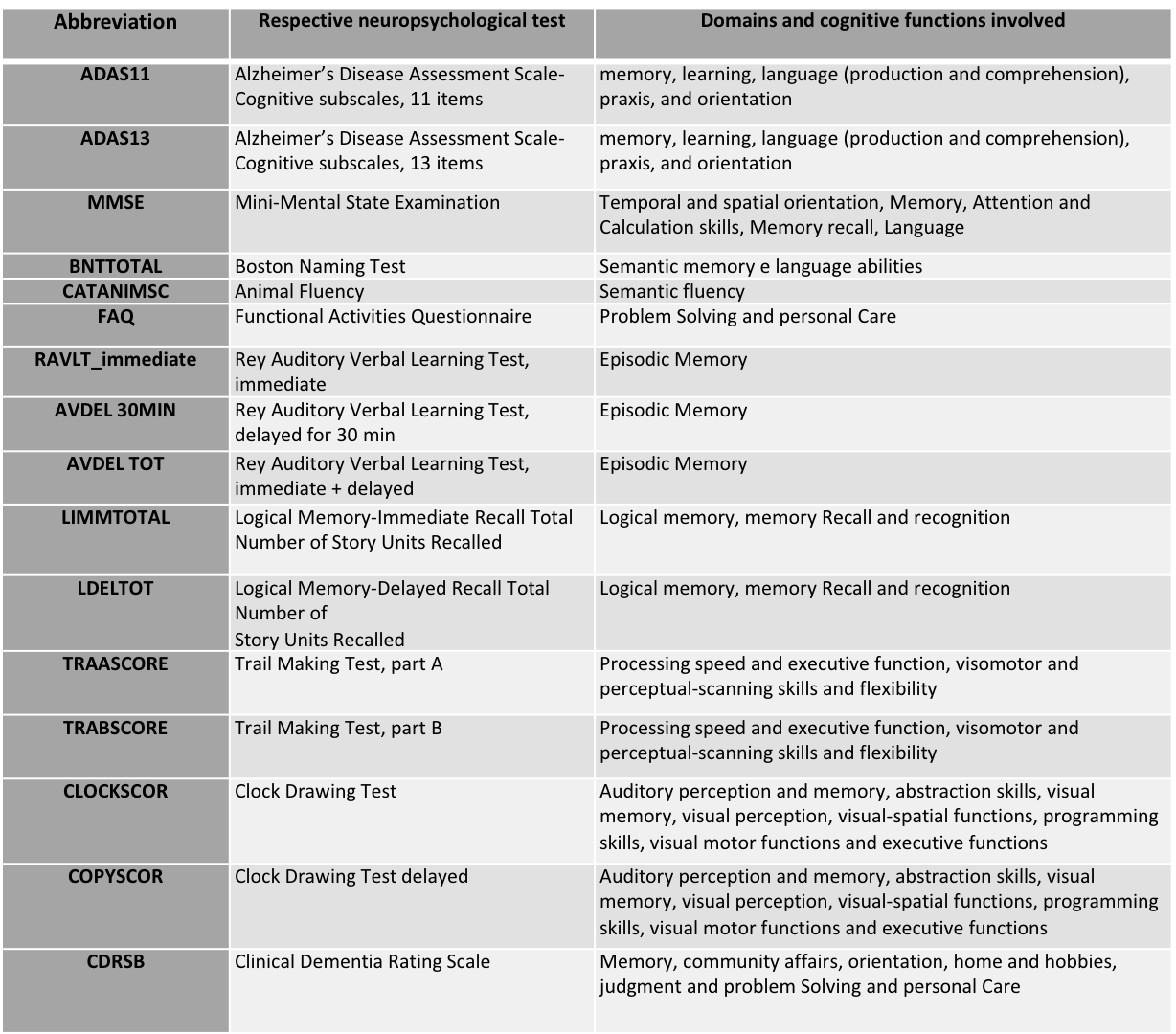
