## Supplementary Table 4 for "A machine learning-based holistic approach for diagnoses within the Alzheimer’s disease spectrum"

**Supplementary Table 4. List of all the evaluated biomarkers and the biospecimens from which they were extracted**.

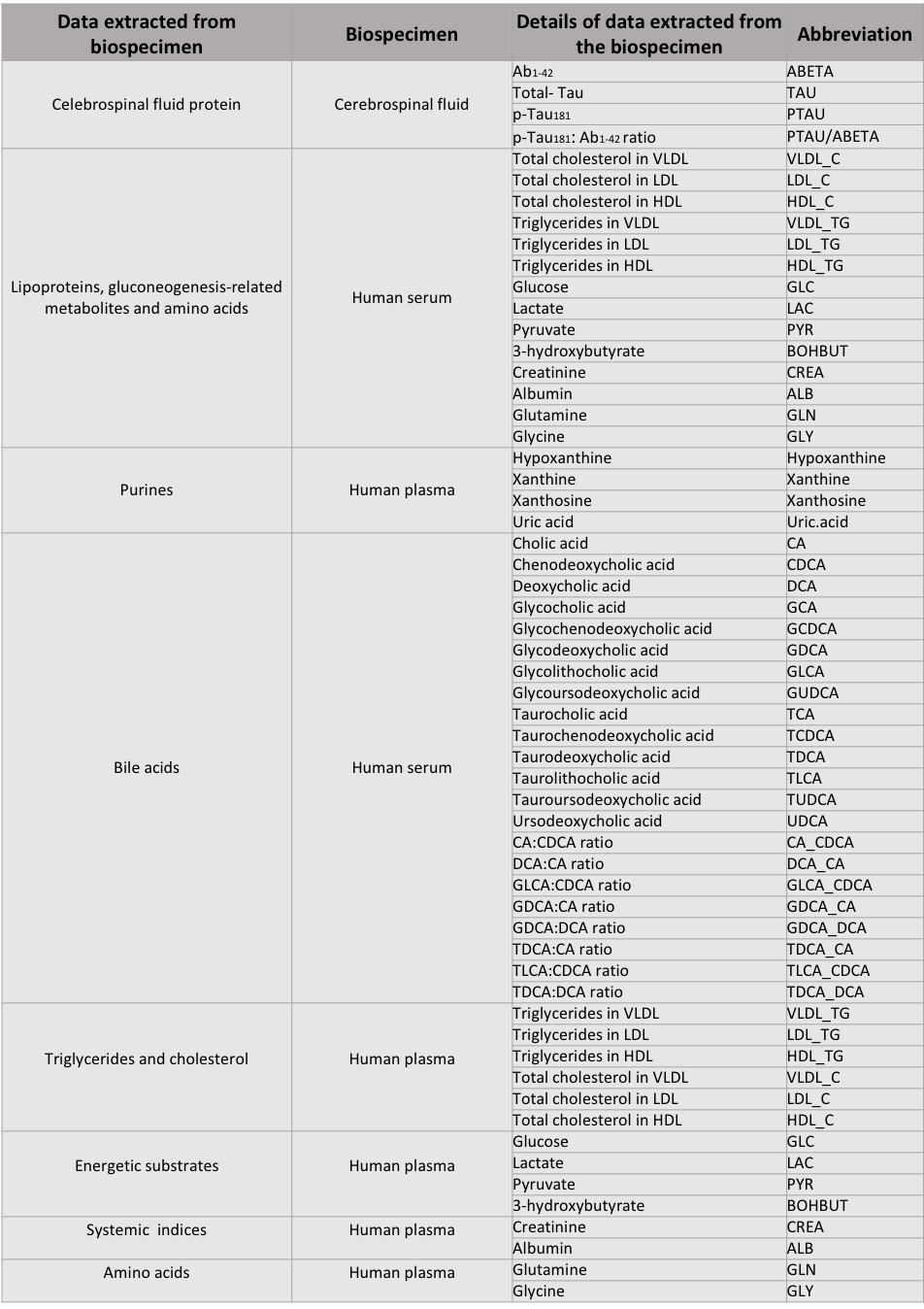

CSF protein levels were measured using the Roche fully automated immunoassay platform (Cobas e601) and immunoassay reagents. The quantifications of lipoproteins and various low-molecular metabolites (including amino acids) were obtained using the Nightingale Health's metabolomics platform, whose functioning is based on the combination of Nuclear Magnetic Resonance and mass spectroscopy. The sample analysis of bile acids was performed by ultra-high pressure liquid chromatography tandem mass spectrometry with the Biocrayes Bile Acids assay. The AD-related metabolomic data collected by Alzheimer's Disease Metabolomics Consortium (ADMC) were obtained by mass spectroscopy using targeted and non-targeted metabolomics platforms. Finally, purine quantifications were analyzed by liquid chromatography (Agilent Technologies 1200) coupled to electrospray ionization on a triple quadrupole mass spectrometer (Agilent Technologies).
