## Supplementary Table 5 for "A machine learning-based holistic approach for diagnoses within the Alzheimer’s disease spectrum"

**Supplementary Table 5. List of brain areas considered according to the Desikan-Killiany Atlas nomenclature.**

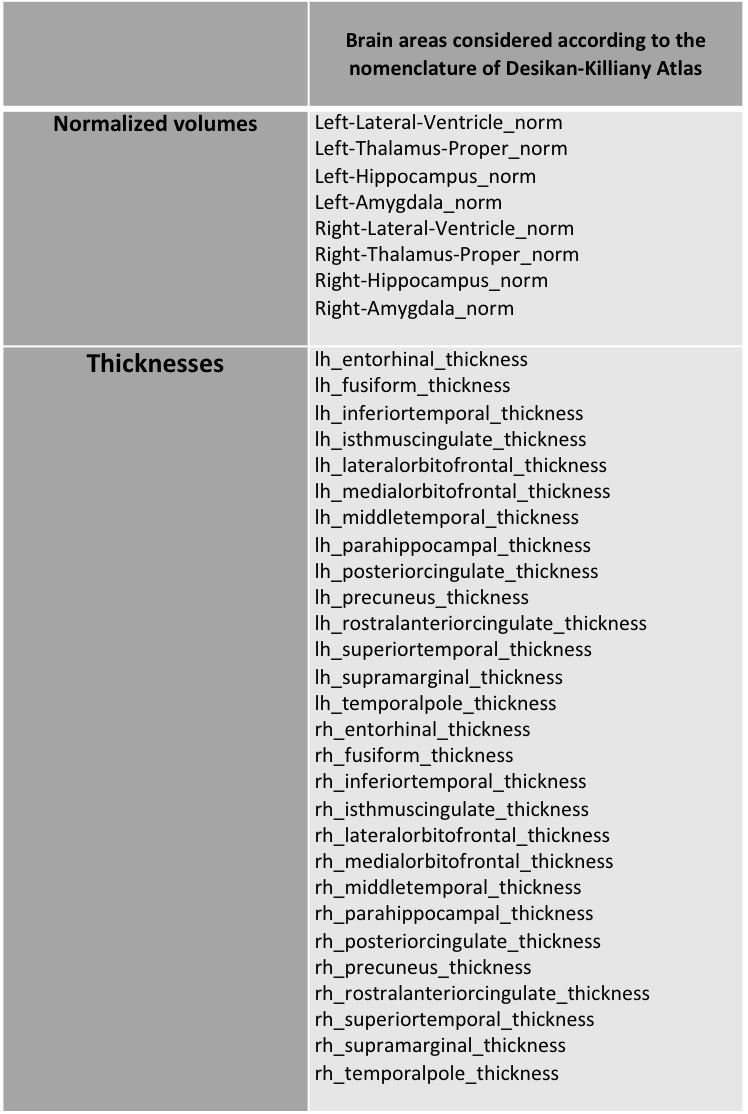
