## Supplementary Figure 1 for "A machine learning-based holistic approach for diagnoses within the Alzheimer’s disease spectrum"

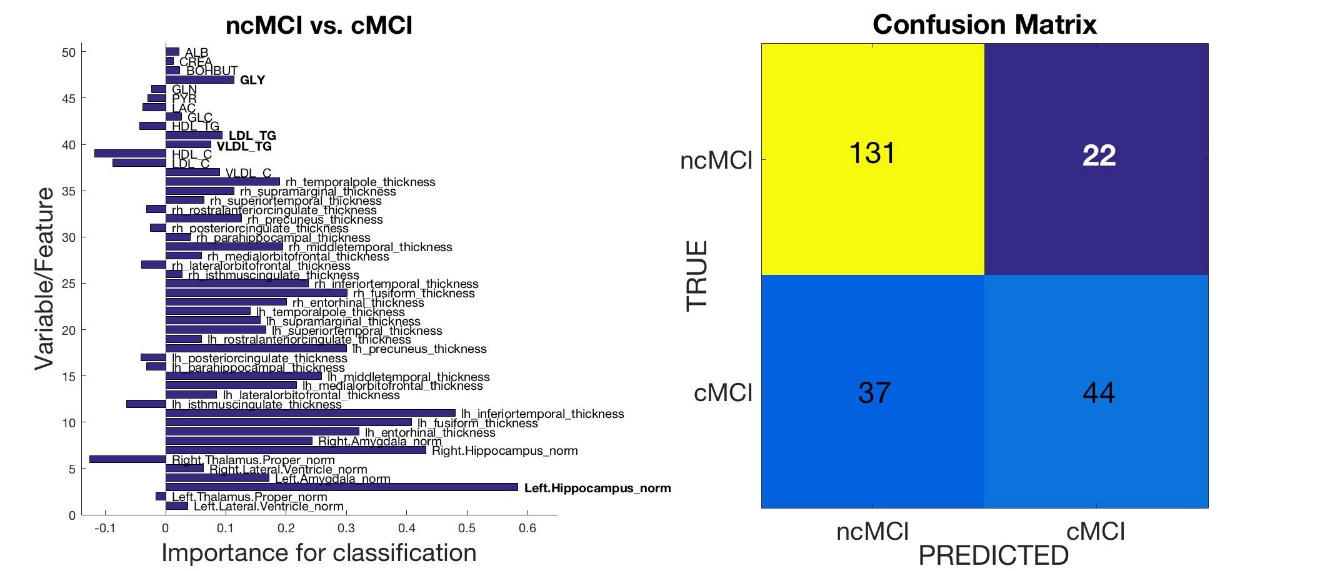

**ML training phase output for old (> 70 years old) MCI subjects**. The figure illustrates the outcome of the RF analysis using as feeding data the metabolic and structural MRI data. (Left) The histogram depicts the classification weight, or importance, of each feature employed for predicting conversion (cMCI) or non-conversion (ncMCI) of old (> 70 years old) MCI subject to AD. Classification scores range from 0 to 1, with higher values indicating better classification abilities. Note that the primary drivers for classification and prediction of ncMCI subjects are the values of left hippocampal volumes and metabolic indices (i.e.: lipoproteins and glucogenesis-associated metabolites; see also Supplementary Table 4 for a full list of the evaluated features). (Right) The pictogram illustrates the supervised ML Confusion Matrix. The Matrix provides an overview of the ML ability to predict conversion (cMCI) or non-conversion (ncMCI) of MCI young subjects to AD. The upper left box shows the number of true positive ncMCI subjects recognized by the ML; the upper right box, instead, shows the number of false positive ncMCI subjects identified by the algorithm. The bottom left square shows the number of cMCI individuals identified as ncMCI (false positive) by the ML; the bottom right box shows the number of true positive cMCI subjects recognized by the algorithm.
